# Improvements in Family Member Quality of Life Following a Task-Shifted Psychosocial Intervention for Road Traffic Injury Survivors in Cameroon

**DOI:** 10.64898/2026.08.10.26360156

**Authors:** Arole Darwin Touko, Fanny Nadia Dissak Delon, Gyscard Merlin Pola, Rasheedat Oke, Catherine Juillard, Alain Chichom Mefire, Sithombo Maqungo

## Abstract

**Background:** Road traffic injuries (RTI) -related trauma is associated with heavy psychosocial burdens for family members involved as informal caregivers after the event. Little is known about the quality of life (QoL) experience of family members of RTI survivors in sub-Saharan Africa. This study investigated the QoL outcomes of family members after the administration of the Friendship Bench psychosocial intervention among RTI survivors in Cameroon.

**Methods:** A pre-post study design was utilized in two trauma centers in Cameroon. The relatives underwent assessment both at baseline (Session 1) and follow-up sessions (Session 6) using the WHOQOL-BREF tool, without being exposed to any intervention. The domain scores were scaled from 0 to 100. Statistical analysis consisted in paired t-tests/Wilcoxon signed-rank tests along with effect sizes using Cohen’s d.

**Results:** The study involved twenty-nine participants. Baseline assessment revealed overall poor quality of life across all domains, with scores in the poor range (Social = 33.33 [IQR: 25.00–47.92]; Environmental = 34.38 [IQR: 25.78–45.31]; Psychological = 45.83 [IQR: 34.38–50.00]; Physical = 46.43 [IQR: 39.29–56.25]). These scores had a statistically significant increases across all domains after the intervention (p < 0.001), accompanied by medium to large effect sizes (Cohen’s d:Social = 1.35; Environmental = 1.57; Psychological = 1.46; Physical = 0.73). The relationship type was found to predict social QoL (F(6,22) = 4.57, p = 0.004), with children of patients demonstrating the greatest gains.

**Conclusion:** Family members of RTI survivors showed a significant improvement in QoL during the Friendship Bench intervention. These results highlight the importance of evaluating family member outcomes in task-shifted psychosocial programs in LMICs.

## 1. Introduction

Road traffic injuries (RTIs) represent a major global public health challenge, accounting for approximately 1.19 million deaths annually and constituting the leading cause of death among individuals aged 5–29 years [1]. The burden is disproportionately concentrated in low- and middle-income countries (LMICs), particularly in sub-Saharan Africa, where weak health systems, limited trauma care infrastructure and inadequate emergency services contribute to high mortality and long-term disability rates [1,2]. In Cameroon, these systemic constraints further exacerbate recovery outcomes, leaving many survivors with prolonged physical, psychological and social challenges [3].

Beyond physical injury, RTIs are strongly associated with adverse mental health outcomes, including depression, anxiety and reduced quality of life among survivors [4–7]. However, the impact of trauma extends beyond the individual patient to affect family members and household systems. Following injury, relatives often assume informal caregiving roles, providing physical, emotional and financial support, frequently without training or external assistance [8,9]. This transition can lead to significant caregiver burden, characterized by psychological distress, social disruption and reduced quality of life [8–10].

Existing literature demonstrates that caregiver burden is substantial in trauma contexts, though most evidence originates from high-income settings and focuses on specific conditions such as traumatic brain injury and spinal cord injury [10,11]. In LMICs, these burdens are likely intensified by economic constraints, limited access to rehabilitation services and strong social expectations of family responsibility [12]. Despite this, there remains a notable lack of research specifically examining the quality of life of family members of RTI survivors, particularly in sub-Saharan Africa.

Emerging evidence suggests that patient-centered psychosocial interventions may generate indirect or “spillover” benefits for household members [13,14]. Improvements in patient’s mental health and functional status may reduce caregiving demands and enhance overall household’s well-being. The Friendship Bench intervention, a task-shifted psychological therapy model implemented in several African settings, has demonstrated effectiveness in improving patient mental health outcomes [15,16]. However, its potential impact on family members’ quality of life has not been explored in the context of RTIs.

This study addresses these gaps by prospectively assessing the quality of life of family members of RTI survivors undergoing the Friendship Bench intervention in Cameroon. We hypothesized that improvements in patient well-being may translate into measurable gains in family members’ quality of life, reflecting broader household-level benefits.

## 2. Methods

### 2.1 Study design

This study employed a prospective pre-post design nested within the intervention of the Friendship Bench for RTI survivors conducted at two trauma hospitals in Cameroon. Family members were assessed at two time points; baseline (concurrent with the patient’s first intervention session) and follow-up (concurrent with the patient’s completion of all six intervention sessions), without themselves receiving any intervention. The six sessions were delivered over a period of six to eight weeks after hospital discharge, with the exact duration varying according to patient availability. For clarity, T0 denotes the baseline assessment (Session 1) and T1 denotes the follow-up assessment (Session 6 completion). This design allows for the estimation of within-person change in QoL over the intervention period and exploration of associations between patient recovery and family member well-being.

### 2.2 Study setting

The study was conducted at Hôpital Laquintinie de Douala and at the Hôpital Régional de Bertoua in Cameroon. Cameroon is a lower-middle-income country in Central Africa with a population of approximately 27 million, characterised by significant linguistic, cultural and geographic diversity [24]. The country’s healthcare system is structured into three tiers (primary, secondary, and tertiary), with trauma care predominantly concentrated in district and regional hospitals in urban centres. Road traffic injuries are a leading cause of hospital admission, placing substantial demands on surgical, orthopaedic and emergency services [5]

### 2.3 Study population

The study involved two distinct populations: **Patient population (intervention recipients):** RTI survivors who presented to participating trauma hospitals following road traffic crashes, were enrolled in the hospital trauma registry, met eligibility criteria for the Friendship Bench intervention and provided informed consent to participate upon presentation of the information notice.

**Family member population (study participants)**: Adult household members of enrolled RTI survivors. These individuals were not themselves RTI survivors, did not receive any component of the Friendship Bench intervention, and were enrolled to assess the potential spillover effects of the patient’s intervention on household quality of life.. Each family member was paired to the specific patient in their household, and the changes in family member QoL (ΔQoL) were analysed in relation to the changes observed in that patient (ΔPHQ-8). The number of family members per patient varied: some had one family member enrolled, while other had more than one, reflecting the natural composition of each household. All enrolled family members who met the eligibility criteria and provided informed consent were included, regardless of how many were linked to a single patient.

For the purpose of this paper, all analyses and reporting pertain exclusively to the family member population.

### 2.4 Inclusion and Exclusion criteria

Family members were considered eligible for inclusion if they were aged 21 years or older at the time of enrolment and resided in the same household as the enrolled RTI survivor. Residence within the same household was defined as sharing a living space and regularly participating in the domestic economy of the household. Eligible participants were required to be able to provide verbal informed consent and to be available and willing to participate in both baseline and follow-up assessments. Additionally, participants were required to be able to communicate in English or French.

Family members were excluded from participation if they were themselves survivors of a current or recent (within the preceding 12 months) road traffic injury. Individuals who were currently enrolled in a formal mental health treatment programme were not eligible for inclusion. Finally, participants who did not reside in the same household as the RTI survivor throughout the study period, including those who relocated or were hospitalized for a prolonged duration, were excluded from the study

### 2.5 The Friendship Bench Intervention

The Friendship Bench is a structured, task-shifted psychological intervention developed by Professor Dixon Chibanda and colleagues in Zimbabwe and subsequently adapted in several sub-Saharan African countries [22,23]. The intervention is delivered by trained lay health workers often referred to as “Grandmothers” or community health workers who are supervised by mental health professionals. It integrates principles of problem-solving therapy (PST) with culturally adapted counselling techniques designed to address common mental disorders including depression, anxiety, and psychosocial distress.

Sessions were delivered by trained research assistants with no prior mental health background or clinical experience, consistent with the task-shifting principles underpinning the Friendship Bench model. Research assistants underwent a three-day structured training programme, which included didactic instruction, role-play, and supervised practice sessions. Weekly supervision was provided throughout data collection, including group feedback sessions to monitor fidelity and address challenges in delivery. Sessions lasted between 45 to 60 minutes each. Sessions included: (1) rapport building and psychoeducation; (2) problems and goals identification; (3) problem-solving techniques; (4) making decisions and planning actions; (5) implementing actions and reviewing their results; and (6) consolidating. All sessions were delivered in person, no remote or telephone delivery was used.

In the current Cameroonian adaptation, the intervention was a 6-8 weeks procedure comprising six structured sessions delivered after patient’s discharge from the hospital.. Each session addressed specific domains of psychological distress, coping and problem-solving capacity, with progressive skill-building across sessions. Sessions contents included psychoeducation about mental health and injury recovery, identification of personal strengths and resources, structured problem-solving exercises and enhancement of social support networks.

### 2.6 Data Collection Procedures

RTI survivors were approached for enrolment during hospital admission. Following patient enrolment, household members were subsequently recruited during the first home visit conducted for the initial session of the Friendship Bench intervention. At this visit, all family members present in the household who met the eligibility criteria were invited to participate in the study. There was no predetermined limit on the number of participants per household; therefore, all available and eligible household members who consented were enrolled.

#### 2.6.1 Baseline assessment

The baseline assessment was conducted at the time of the patient’s first Friendship Bench session (Session 1). The family member was interviewed separately from the patient to prevent social desirability bias and ensure independent response. Interviews were conducted in the respondent’s preferred official language (French or English).

At baseline, family members completed: 1)-a sociodemographic questionnaire (age, sex, relationship to patient, educational attainment, occupation, household income, number of household members, duration of caregiving since injury) and 2)-the WHOQOL-BREF checklist.

#### 2.6.2 Follow-up assessment

The follow-up assessment was conducted after the RTI survivor completed all six Friendship Bench sessions (week 6 or 8). We administered the WHOQOL-BREF and collected brief information on any significant life events or changes in household circumstances since baseline.

### 2.7 Outcome measures

The primary outcome was quality of life as measured by the WHOQOL-BREF [25,26]. The WHOQOL-BREF was selected because of its cross-cultural validity, its demonstrated applicability in LMIC settings and its ability to capture multiple dimensions of well-being relevant to the experience of family members of trauma survivors. The WHOQOL-BREF comprises 26 items assessed on a five-point Likert scale

### 2.8 Statistical Analysis

The analyses were performed in R version 4.3. Results of descriptive statistics were presented as median [interquartile range (IQR)] throughout, and categorical variables as frequency and percentage. Given the small sample size, the median was chosen as the primary mesure of central tendency regardless of distribution. The normality of the change score distribution was tested using the Shapiro-Wilk test. Paired sample t-tests were performed for the normally distributed change scores; otherwise, the Wilcoxon signed-rank test was conducted. Effect sizes were measured as Cohen’s d for paired samples, defined as the mean difference divided by the standard deviation of individual differences. Multiple linear regressions were carried out to find out sociodemographic predictors of change in the respective QoL domains. Spearman’s rank correlation was estimated to explore the relationship between the change in PHQ-8 score among patients (ΔPHQ-8) and the difference in QoL (ΔQoL) among their families. Differences in change in each QoL domain according to relationship type were tested using one-way ANOVA with Tukey HSD post hoc tests. P < 0.05 was considered statistically significant; 95% confidence intervals (CIs) were given for the primary outcomes.

### 2.9 Ethical consideration

This study was conducted in accordance with established ethical standards. Ethical approval was obtained from the University of Cape Town Human Research Committee (HREC REF: 611/2025) and the University of Buea Institutional Review Board (2025: 2003-04/UB/SG/IRB/FHS). Administrative authorization was obtained from the Regional Delegations of Public Health for the Littoral and East regions under authorization numbers 403/AR/MINSANTE/SG/DRSPE/BCASS/C2/CA and 0168/AR/MINSANTE/DRSPL/BCASS respectively. Verbal informed consent was obtained from all participants after presentation of the information sheet.

## 3. Results

### 3.1 Sample characteristics and data quality

Twenty-nine family member-patient’s with complete data at both time points were included in the analysis (BRH: n=14; HLD: n=15).

### 3.2 Sociodemographic Characteristics

The median age of participants was 29 [IQR: 25–47] years (BRH: 28 [IQR: 25–35]; HLD:30 [IQR: 24–56]. Females predominated (69.0%) in the overall sample, with higher predominance at BRH (78.6%) compared to HLD (60.0%). Most participants had secondary education (51.7%) and those who had university were 34.5%. Most of the participants were single (41.4%), and those at BRH were 42.9%,. Cohabitation was more common at BRH (50.0%) than at HLD (0.0%). Self-employed participants were 27.6%, and unemployed participants were 20.7%. The category of siblings was the highest (24.1%), and those of children and spouses were 20.7%. HLD had higher numbers of children of patients (33.3%) compared to BRH (7.1%), and BRH has higher numbers of spouses (28.6%) and uncle/aunty (21.4%). Full characteristics are presented in Table 1.

**Table 1:**
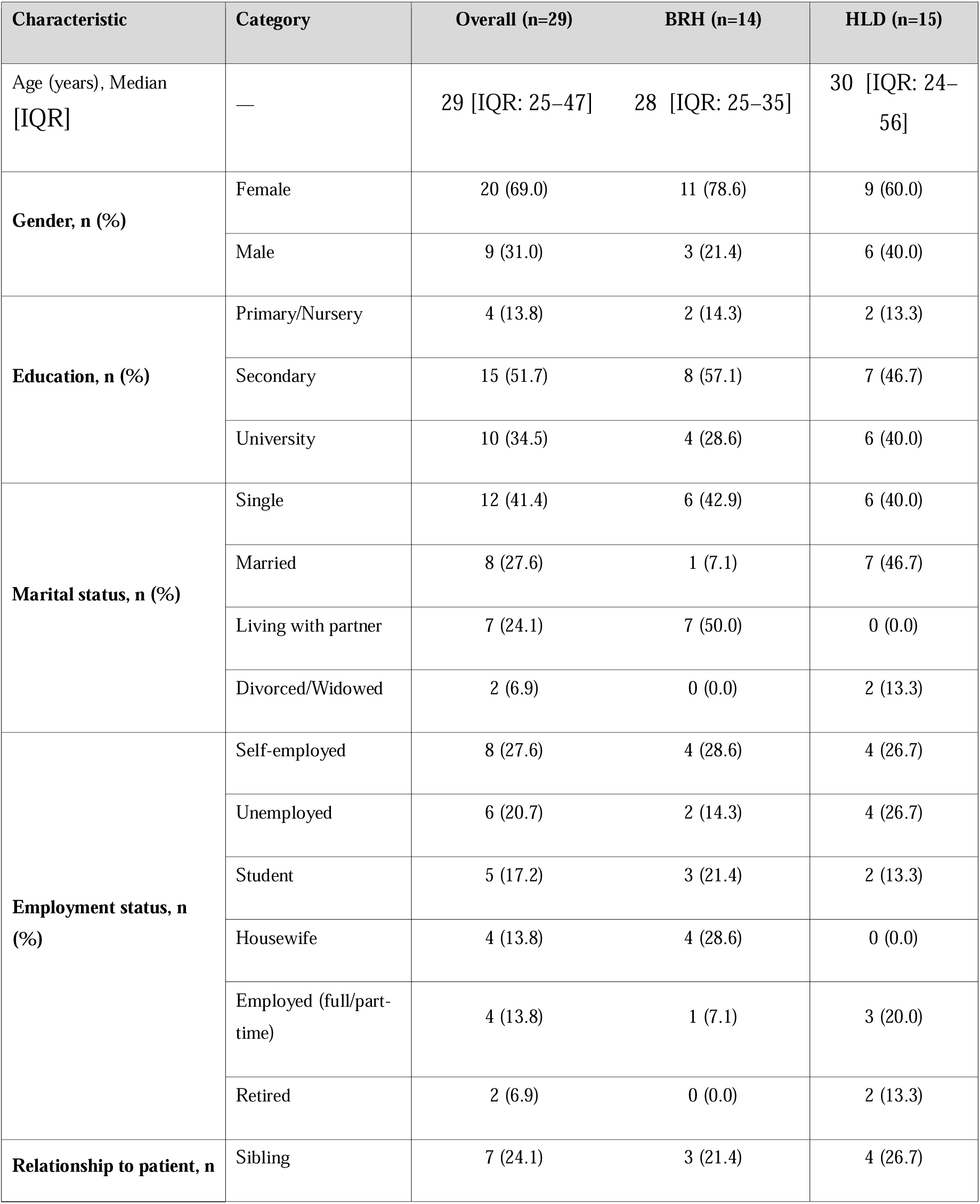
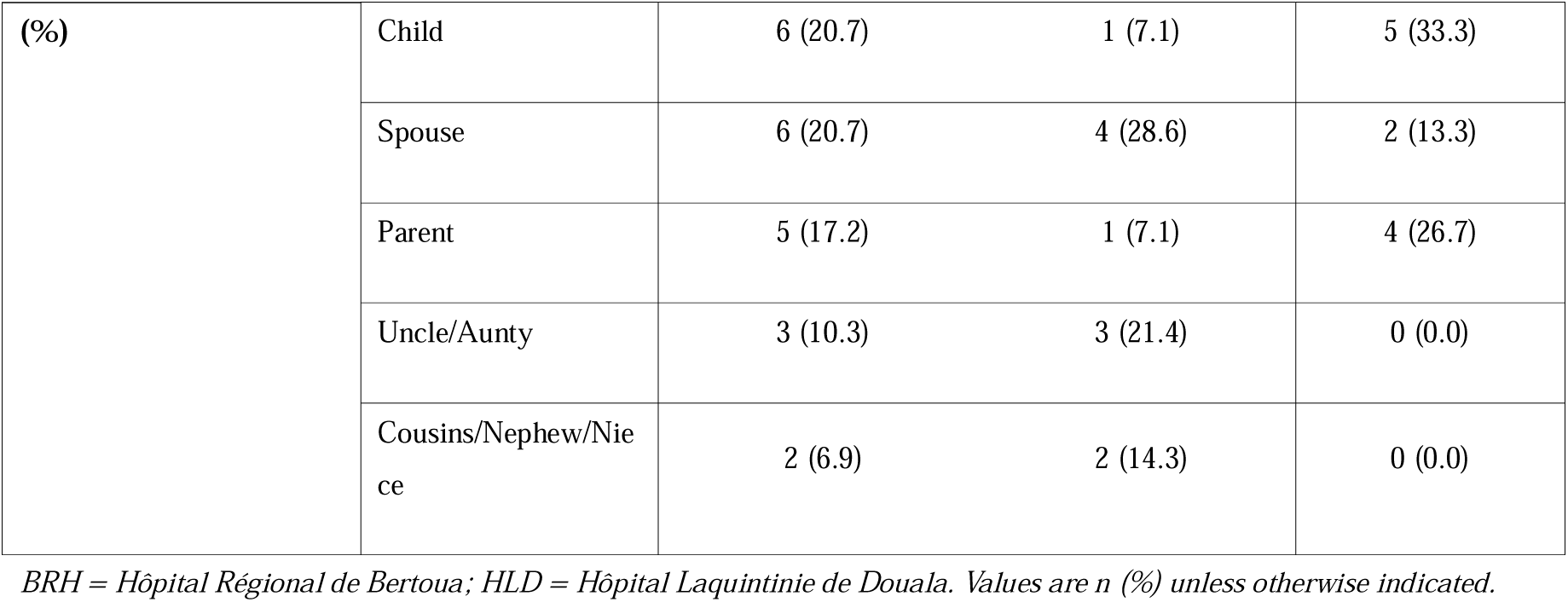
Baseline sociodemographic characteristics of participants.

### 3.3 WHOQOL-BREF Domain Scores at Baseline (T0)

At baseline, the four domains of the WHOQOL-BREF questionnaire and the general QoL had their values in the poor quality of life category (values between 25–49 according to the transformed 0-100 scale). The social domain recorded the lowest median score (33.33 [IQR: 25.00–47.92]), with considerable heterogeneity across participants (range: 0.00–91.67). The environmental domain was similarly low (median: 34.38 [IQR: 25.78–45.31]). The psychological (median: 45.83 [IQR: 34.38–50.00]) and physical (median: 46.43 [IQR: 39.29– 56.25]) domains recorded the highest values at baseline, though both remained within the poor QoL range. General QoL had a median of 37.50 [IQR: 25.00–50.00], with values ranging from 0.00 to 87.50, indicating substantial variability in participants’ overall perceived quality of life at the time of the patient’s first intervention session. Baseline descriptive data is provided in Table 2, while Figure 1 shows the difference between T0 and T1 in all domains.

**Figure 1:**
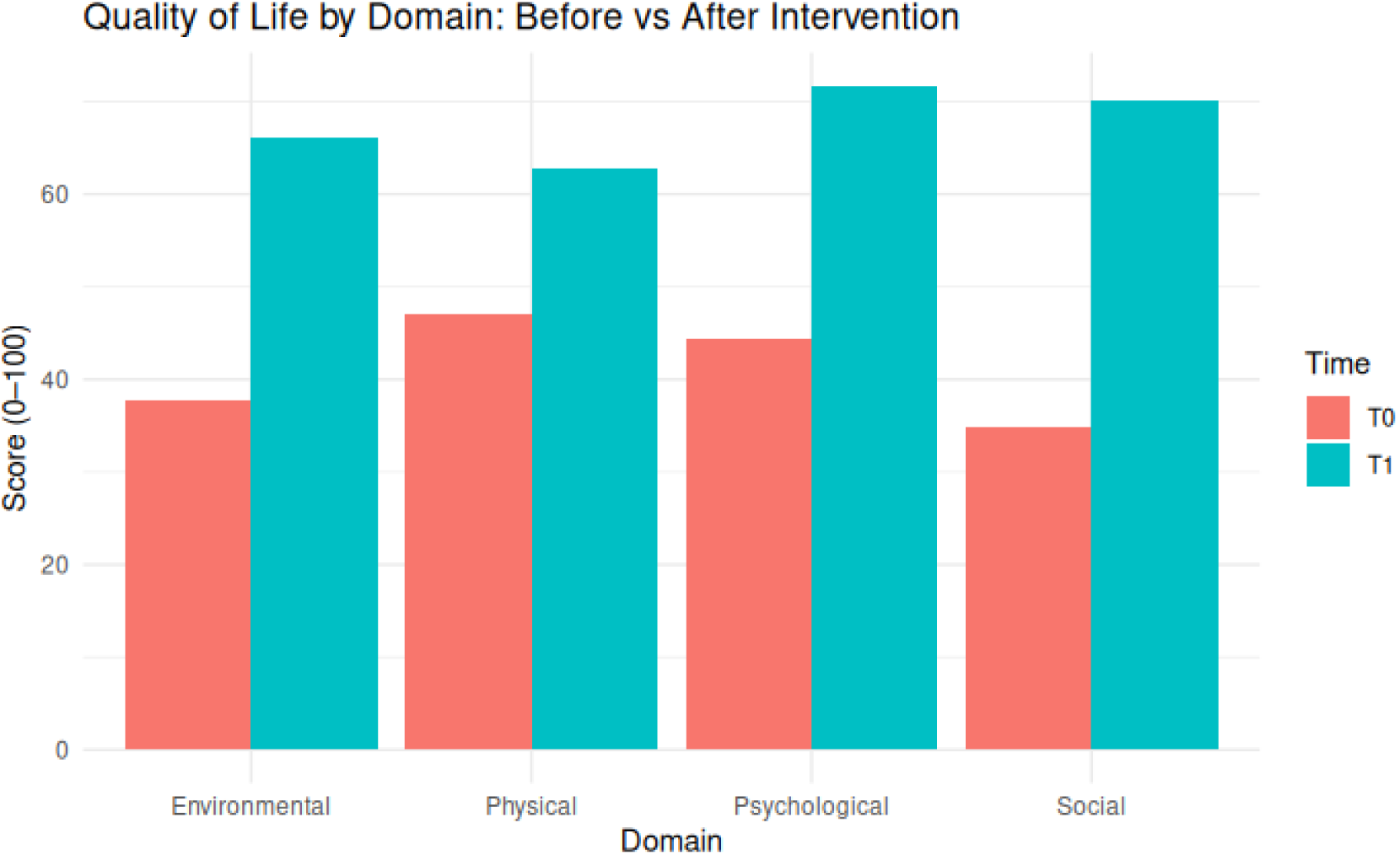
Quality of life by Domain: Before vs After Intervention

**Table 2:** Descriptive statistics of WHOQOL-BREF domain scores at baseline (T0)

| Domain | Min | Q1 | Median | Mean | Q3 | Max |
| --- | --- | --- | --- | --- | --- | --- |
| Physical | 3.57 | 39.29 | 46.43 | 47.02 | 56.25 | 75.00 |
| Psychological | 8.33 | 34.38 | 45.83 | 44.31 | 50.00 | 83.33 |
| Social | 0.00 | 25.00 | 33.33 | 34.84 | 47.92 | 91.67 |
| Environmental | 12.50 | 25.78 | 34.38 | 37.81 | 45.31 | 81.25 |
| General QoL* | 0.00 | 25.00 | 37.50 | 39.17 | 50.00 | 87.50 |

*\*General QoL represents the composite of Items 1 and 2 of the WHOQOL-BREF, scored on a 0–100 scale and reported separately from the four primary domains. Q1 = first quartile; Q3 = third quartile.*

### 3.4 WHOQOL-BREF Domain Scores at Follow-up (T1)

We observed a significant change at the follow-up stage (week 6-8), as all domains of WHOQOL-BREF moved into the range of moderate to good quality of life (scores 50-74). The psychological domain recorded the highest median score (70.83 [IQR: 62.50–83.33]), followed by the social (75.00 [IQR: 58.33–83.33]), environmental (68.75 [IQR: 50.00– 75.00]), and physical (60.71 [IQR: 53.57–67.86]) domains. It is noteworthy that while the minimal value of the physical domain was very low, amounting to only 3.57, it rose up to 46.43 at the follow-up stage. At baseline, the social domain received the minimal value of 0.00, and at the second testing moment, it got 25.00 points. The maximal values of psychological, social, and environmental domains amounted to 95.83, 100.00, and 100.00 correspondingly, which shows that some participants scored perfectly well in those domains. General QoL median increased two-fold, rising from 37.50 to 75.00 points at follow-up [IQR: 62.50–75.00].Detailed T1 descriptive statistics are presented in Table 3.

**Table 3:** Descriptive statistics of WHOQOL-BREF domain scores at follow-up (T1).

| Domain | Min | Q1 | Median | Mean | Q3 | Max |
| --- | --- | --- | --- | --- | --- | --- |
| Physical | 46.43 | 53.57 | 60.71 | 62.81 | 67.86 | 89.29 |
| Psychological | 50.00 | 62.50 | 70.83 | 71.70 | 83.33 | 95.83 |
| Social | 25.00 | 58.33 | 75.00 | 70.11 | 83.33 | 100.00 |
| Environmental | 40.62 | 50.00 | 68.75 | 66.06 | 75.00 | 100.00 |
| General QoL* | 37.50 | 62.50 | 75.00 | 71.12 | 75.00 | 100.00 |

\*General QoL represents the composite of Items 1 and 2. Q1 = first quartile; Q3 = third quartile.

### 3.5 Pre-Post Comparison of QOL Domain Scores

Statistically significant improvement among participants was found in all four WHOQOL-BREF domains as well as in overall QoL. Normality of change scores for each domain was proven by using the Shapiro-Wilk test: for Physical (W=0.966; p=0.456), for Psychological (W=0.975; p=0.704), for Social (W=0.977; p=0.762) and for Environmental (W=0.973; p=0.643). Change scores for General QoL were not normally distributed (W=0.911; p=0.018), thus a Wilcoxon signed-rank test was needed.

A statistically significant improvement was achieved for the physical domain (mean difference = 16.01, 95% CI: 7.65–24.37; t = 3.92, df = 28, p < 0.001; Cohen’s d = 0.73). For the psychological domain, there was a very large improvement that proved to be highly significant (mean difference = 27.44, 95% CI: 20.30–34.59; t = 7.87, df = 28, p < 0.001; Cohen’s d = 1.46). In the social domain, a highly significant improvement was observed (mean difference = 34.36, 95% CI: 24.70–44.02; t = 7.29, df = 28, p < 0.001; Cohen’s d = 1.35). In addition, an increase in the environmental domain was highly significant as well and amounted to a very large improvement (mean difference = 28.02, 95% CI: 21.21–34.82; t = 8.44, df = 28, p < 0.001; Cohen’s d = 1.57). General QoL increased significantly according to the Wilcoxon signed-rank test. All results are shown in Table 4.

**Table 4:** Comparison of WHOQOL-BREF domain scores between baseline (T0) and follow-up (T1)

| Domain | T0 Mean ± SD | T1 Mean ± SD | Mean Diff (95% CI) | Statistics | p-value | Cohen's d |
| --- | --- | --- | --- | --- | --- | --- |
| Physical | 46.80 ± 16.49 | 62.81 ± 11.31 | 16.01 (7.65–24.37) | $t = 3.92$ | $< 0.001$ | 0.73 |
| Psychological | 44.25 ± 15.45 | 71.70 ± 12.76 | 27.44 (20.30–34.59) | $t = 7.87$ | $< 0.001$ | 1.46 |
| Social | 35.75 ± 22.10 | 70.11 ± 20.48 | 34.36 (24.70–44.02) | $t = 7.29$ | $< 0.001$ | 1.35 |
| Environmental | 38.04 ± 17.97 | 66.06 ± 16.42 | 28.02 (21.21–34.82) | $t = 8.44$ | $< 0.001$ | 1.57 |
| General QoL | 39.17 (IQR: 25.0–50.0) | 75.00 (IQR: 62.5–75.0) | — | $V = 253$ | $< 0.001$ | 1.15 |

### 3.6 Ranking of Domain by Absolute and Relative Improvement

With respect to improvement, in absolute terms, the social dimension was the highest, scoring an improvement of +33.33 points, followed by environmental (+28.13), psychological (+25.00), general (+25.00), and physical (+10.71). With respect to percentage increase in relation to initial values, the social domain again had the largest increase, having improved by 125.0%, followed by environmental (100.0%), general (100.00%), psychological (54.55%), and physical (30.77%) domains, respectively. It is interesting to note that the dimensions with the lowest initial values, which are also those with the greatest needs, experienced the highest percentage improvement. The ranking can be observed in Table 5.

**Table 5:** Ranking of WHOQOL-BREF domains by absolute and percentage improvement from T0 to T1.

| Domain | T0 Median | Median difference | % Improvement |
| --- | --- | --- | --- |
| Social | 33.33 | 33.33 | 125.00% |
| General QoL | 37.50 | 25.00 | 100.00 % |
| Environmental | 34.38 | 28.13 | 100.00% |
| Psychological | 45.83 | 25.00 | 54.55 % |
| Physical | 46.43 | 10.71 | 30.77% |

*Percentage improvement = (T1 Median – T0 Median) / T0 Median × 100. Domains ranked in descending order of absolute improvement.*

### 3.7 Predictors of Change in Quality of Life Domains

Multivariate regression was performed separately for each domain of quality of life based on Δ=T1-T0 scores as a dependent variable, and the independent variables included age, gender, education and relationship type. Out of five regression models, only that for social quality of life was statistically significant (F(10,18)=2.52, p=0.043, R²=0.583, adjusted R²=0.351), whereas the other four models for general (p=0.156), physical (p=0.145), psychological (p=0.895) and environmental (p=0.535) domains of quality of life were not statistically significant.

In the context of the significant model for social quality of life, the type of relationship turned out to be the main predictor. When child of the patient was used as a baseline for comparing all other types of relationship, the parents (β=-58.05, p=0.006) and uncles & aunts (β=-58.68, p=0.009) had significantly lower social quality of life changes in comparison to children. In addition, the siblings (β=-22.55, p=0.085) and spouses (β=-26.45, p=0.086) showed non-statistically significant but borderline trends towards having less improvements of social quality of life in comparison to children. Age, gender, education were not found to be significant in any of the models.

### 3.8 Effect of Relationship Type on Social QoL Change

One way ANOVA indicated there was a significant influence of the relationship type on changes in social quality of life (F(6,22) = 4.57, p = 0.004). For none of the other domains, the relationship type influenced the changes significantly (See Table 7). The post-hoc analysis using Tukey HSD test revealed only two significant pair-wise differences in the mean change scores for social QoL among the relationship types: children versus parents (difference = −50.00, p = 0.004) and children versus uncles/aunts (difference = −50.00, p = 0.018). There were no other pair-wise significant differences. Figure 3 below depicts the results visually using boxplot.

**Figure 2:**
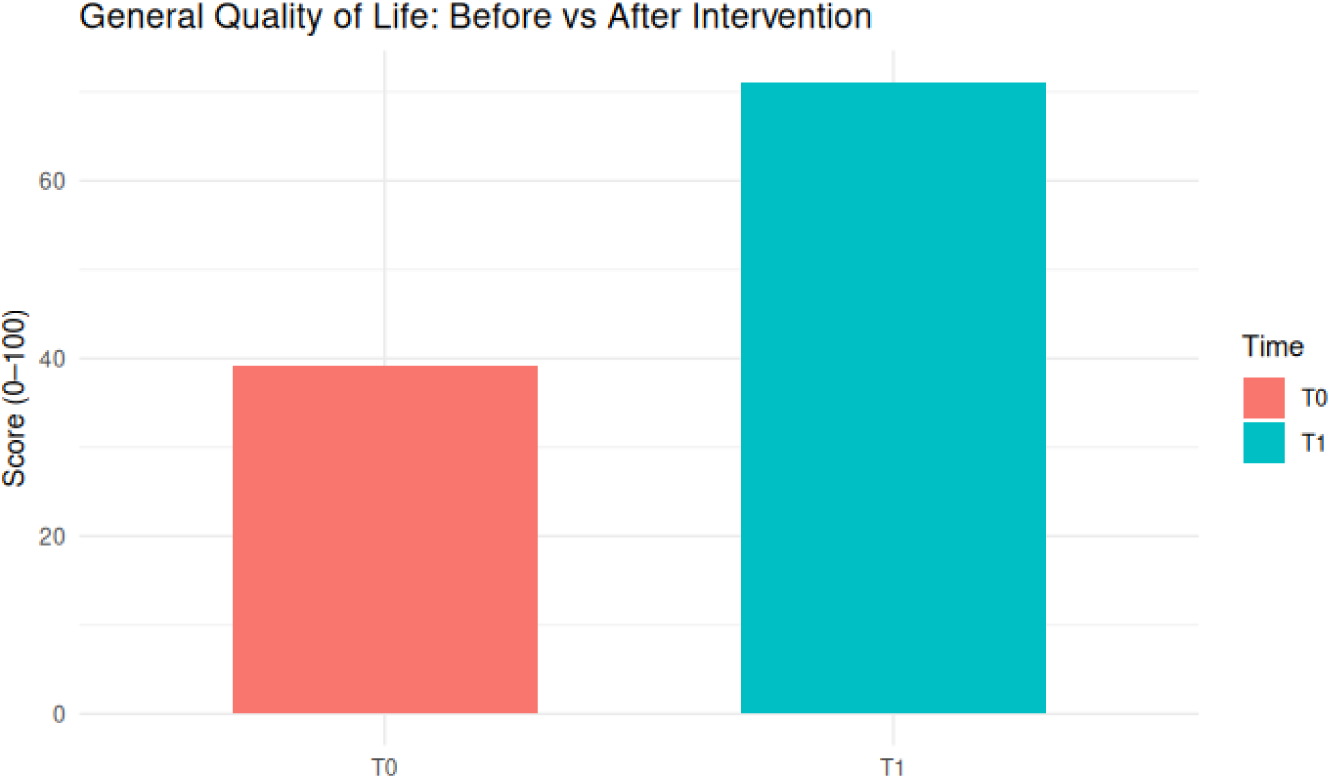
General Quality of Life: Before vs After Intervention

**Figure 3:**
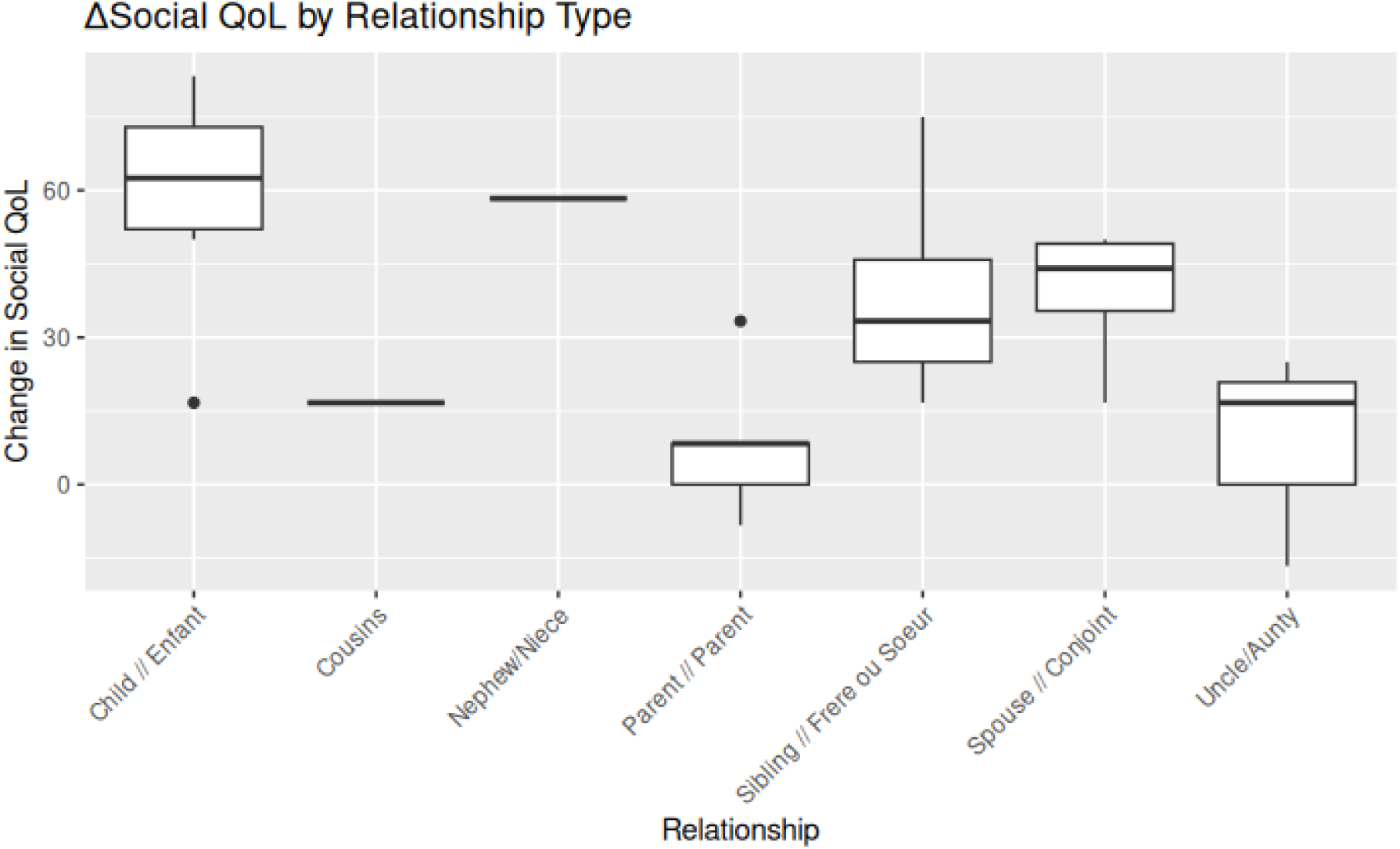
ΔSocial QoL by Relationship Type

**Table 6:**
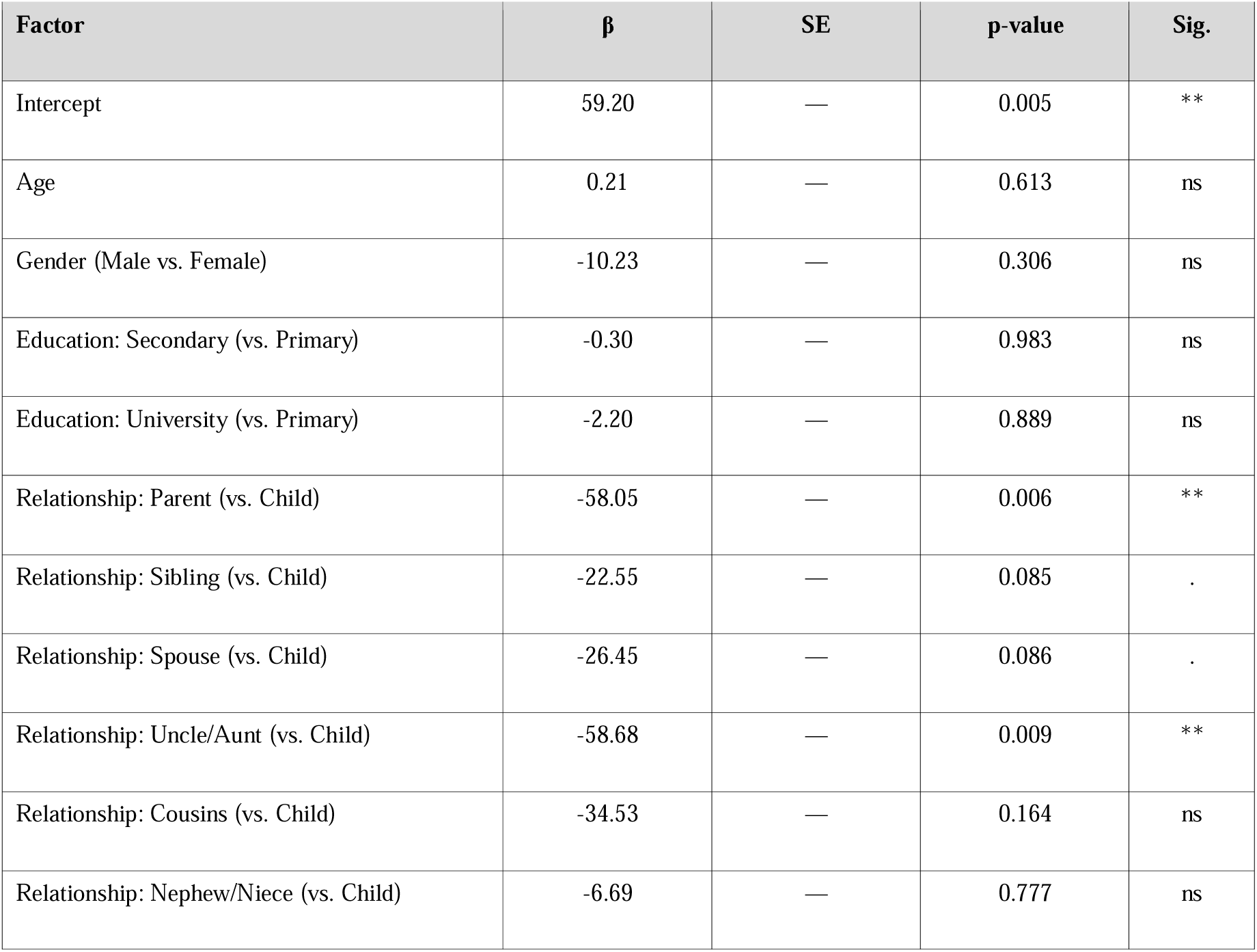
Multivariable linear regression for independent factors associated with change in social quality of life (ΔSocial)

**Table 7:** Spearman rank correlations between ΔPHQ-8 and ΔQOL domain score.

| QoL Domain | Spearman $\rho$ | Direction | Interpretation |
| --- | --- | --- | --- |
| General | -0.208 | Negative | Weak |
| Physical | -0.252 | Negative | Weak |
| Psychological | -0.278 | Negative | Weak |
| Social | -0.023 | Negative | Negligible |
| Environmental | -0.032 | Negative | Negligible |

$\Delta PHQ-8 = PHQ-8 \text{ T1} - PHQ-8 \text{ T0}$ ; negative values indicate improvement. $\Delta QoL = WHOQOL-BREF \text{ domain score T1} - T0$ .

### 3.9 Correlation Between ***Δ***PHQ-8 and ***Δ***QOL

The Spearman’s rank correlation coefficient was applied to a subgroup of 12 patient-family. The Shapiro-Wilk test revealed that ΔPHQ-8 (W = 0.806, p < 0.001) and ΔGeneral QoL (W = 0.912, p = 0.030) were not normally distributed. The correlations between ΔPHQ-8 and ΔQoL for each category using a nonparametric test were low and negative: ΔGeneral (ρ = −0.208); ΔPhysical (ρ = −0.252); ΔPsychological (ρ = −0.278); ΔSocial (ρ = −0.023); and ΔEnvironmental (ρ = −0.032). The inverse relationship can be explained by the scoring criteria: decrease in ΔPHQ-8 implies improvement in QoL. None of these correlations were statistically significant (Table 7).

## 4. Discussion

This study investigated quality of life among relatives of RTI survivors enrolled in the Friendship Bench intervention program in Cameroon, with the objective being to determine whether the intervention can have spillover effects on family members without any form of therapy. The main finding of the study was that there was a significant and very large increase in the quality of life (QoL) among family members in all the five domains of the WHOQOL-BREF Tool without them undergoing the same treatment program.

### 4.1 Baseline Quality of Life: A Portrait of Compromised Wellbeing

The initial scores across all domains of WHOQOL-BREF ranged between poor values (25-49), with the social domain having the lowest median score of 33.33 [IQR: 25.00–47.92]. These trends are supported by literature that examines the impact of caregiving in cases of traumatic events internationally. For instance, studies conducted by Pozzato et al. have established that family members of individuals injured due to accidents on roads have long-term decreases in health-related quality of life, with the social functioning domain being particularly affected [9]. According to Marshall et al., social isolation characterizes TBI caregiver intervention experiences since they often withdraw from social activities in order to fulfill their caregiving duties [10].

Baseline scores for the environmental domain were also quite low 34.38 [IQR: 25.78–45.31], implying that the families considered themselves lacking adequate finances, service provision, safety, and living conditions. It is reasonable to assume that the situation in Cameroon would lead to result in such results: the negative financial impact from the RTI – involving not only medical expenses but also loss of income from working patients and higher expenses at home – could be expected to directly contribute to low resource adequacy perception [12]. Sodders et al. demonstrated that financial strain was an important mediator in the relationship between caregiver burden and quality of life in economically constrained settings [11].

The most variable results were observed for the general QoL (median 37.50 [IQR: 25.00– 50.00]), with the minimum value being equal to 0.00. It can be assumed that a certain percentage of respondents at the moment of the first patient’s intervention experienced poor quality of their lives, implying that some additional help from external sources apart from patient-oriented treatment might be necessary.

### 4.2 Post-Intervention Improvements: Magnitude, Consistency and Clinical Meaning

The positive changes that occurred after the intervention were in line with each other and also were quite significant. All four main areas moved from the poor category to the moderate-to-good category (50-74), and the median score for overall QoL increased twice, from 37.50 to 75.00. The effect size was found to be moderate-to-large (d=0.73 for physical) up to very large (d=1.57 for environmental) with all values exceeding significantly the critical difference of 10 points on the 0-100 scale [17,18].

Effect sizes in the current study are higher than those usually found in interventions involving direct care-giver work in the relevant literature. According to a meta-analysis conducted by Milandeep et al., of psychosocial interventions used with post-traumatic limb amputation patients, effect sizes were moderate (d ≈ 0.5-0.7) for directly treated individuals [13]. The percentages of improvement provide additional perspective to these results. The percentage of improvement was 125.0% in the social domain, 100.0% in general QoL, 100.0% in the environmental domain, 54.55% in the psychological domain, and 30.77% in the physical domain. As the degree of impairment is directly correlated with the percentage of improvement such that the more compromised the domains are, the higher percentages of improvement there were, it can be interpreted as having reached a ceiling effect, which supports the theory of greater potential for improvement.

### 4.3 Domain Specific Findings

The social domain had the lowest score in the pre-intervention phase but finally showed the greatest positive gain (33.33 points). This result can be seen theoretically justified. The Friendship Bench program is based on social interaction; it is administered in a domestic setting by people deeply rooted in the community and focuses on social support as well as addressing interpersonal problems [15,16]. It might be assumed that conducting an intervention within one’s house, even if it concerns oneself, decreases isolation in a household and helps strengthen perceptions of social support within a family. According to Chibanda et al., the social functionality of directly targeted participants also showed positive results in the context of a randomised clinical trial of the program [16].

The environmental domain demonstrated the highest effect size (d = 1.57), although no actual improvement in the participants’ living conditions occurred. The explanation for this result can be provided using cognitive appraisal theory. Since psychological factors play an important role in determining individual perceptions of environmental factors, as one’s emotional and psychological wellbeing improves during the six sessions, he/she may subjectively appraise the quality of his/her environment differently [13]. This statement receives support from the fact that there is a high positive correlation between the scores on psychological and environmental domains, which is a proven characteristic of the WHOQOL-BREF measure [17,18]. In addition, it has already been shown by Pozzato et al. that improved psychological wellbeing after RTI positively influenced the quality of environment scores independently of any actual improvements [9].

Psychological well-being improvements (mean difference [MD] = 27.44, d = 1.46) are one of the most significant results of this study. Family members who provide care to patients after suffering from RTI are at high risk of suffering from depression and anxiety and psychological well-being plays a key role in the overall performance of caregivers [9–11]. The significant improvement in psychological well-being observed in the absence of psychological interventions indicates that the reduction of stress associated with caregiving due to improvement in the condition of patients together with the positive environment created by Friendship Bench meetings might have great psychological effects. According to the theory of spillover effects, the improvement of patient’s condition leads to the reduction of emotional burden of caregiving and redirects psychological efforts toward personal well-being [13,19].

The physical domain had the lowest improvement (mean difference = 16.01, d = 0.73). Health outcomes are not as immediate to changes in psychosocial status within the home setting compared to social and environmental appraisals[10]. Moreover, the pathway by which an intervention would positively affect the non-recipients physically through psychosocial methods would be somewhat indirect, mostly via physical relief from exhaustion and better sleep due to less stress in the home environment. However, despite being relatively small, the effect size of d = 0.73 remains a moderate-to-strong effect size.

### 4.4 Role of Relationship Type in Social QoL Change

The impact of relationship type on social QoL change (F(6,22) = 4.57, p = 0.004) represents a noteworthy additional finding. The children of the patients experienced the largest changes, while the parents and uncles/aunts exhibited notably smaller changes (β = −58.05, β = −58.68 respectively, both p < 0.01). These varying responses have not previously been found in other research on spillover effects in LMICs.

Adult children might gain the greatest advantages from the social contagion effects of their parents’ recovery process because they have a much broader range of social contacts, including their peer groups and work environments, which can be more easily regained as they experience reduced caregiving burdens. However, parents who are caring for injured children might go through a different emotional journey in that their grief and concerns for the wellbeing of their loved ones can prove harder to overcome, even in light of progress in terms of functional recovery, than the patient’s improvement within the six weeks of intervention [10].

The finding that uncles and aunts had shown similar improvements in social QoL as compared to parents can be related to the nature of social dynamics involved when an extended family member serves as a caregiver in the Cameroonian situation, since the extent of obligation towards the wounded members of the extended family is great; however, they do not have the same level of social support that other caregivers such as spousal and filial caregivers do [10]. Marshall et al. indicated that caregivers from an extended family in developing countries tend to suffer more burden due to their lack of closeness to the available resources [10]. The borderline association witnessed among siblings (β = -22.55, p = 0.085) and spouses (β = -26.45, p = 0.086) can be attributed to intermediary dynamics between children and their parents/uncles.

### 4.5 Association Between Patient Recovery and Family QoL

The insignificant and low Spearman’s correlations between changes in PHQ-8 and changes in QoL domains (ρ = −0.023 to −0.278) indicate that the QoL improvements found among family members in the current research have not been directly and significantly related to depression improvements at the level of the individual patients. Instead of a one-directional process where the change in patient depression → reduction in burden of care provided by the patient → family QoL improvements, it can be argued that the latter happen via different and more complicated ways. They can involve changes in communication dynamics within the household, normalization of seeking for help at the family level, establishment of regular routine again, and engagement with the larger community as the household becomes stable throughout the six-sessions period.

### 4.6 Contextualisation Within the Global Literature

This research adds to the developing body of evidence regarding family-level outcomes of psychosocial interventions in LMICs. Studies evaluating the spillover effects of interventions focused on patients have mostly been conducted among people suffering from cancer and chronic disease care-givers in high-income countries [13,14]. Literature related to the Friendship Bench approach, on the other hand, is largely confined to the evaluation of patient-level outcomes [22,23], and family member wellbeing has not been considered much at all. This research addresses this deficiency and shows that community-based psychosocial interventions can yield positive household-level outcomes.

The initial QoL profiles recorded in this study where all aspects fall within the poor category have largely been supported by caregiver studies conducted in similar regions. Hung et al., who investigated the psychological distress among the caregivers of trauma patients in Kenya, found that the caregivers were highly distressed psychologically and socially [2]. Similarly, Tran et al. in their large sample study conducted in Thailand noted that the injured persons’ families continued to suffer from low QoL due to the severe impact of social and psychological domains [14]. It is evident that the initial findings recorded in this study fit well within the regional trend of caregiver experiences within low and middle-income countries within sub-Saharan Africa and South East Asia.

The size of change seen in this study, specifically in the social (d = 1.35) and environmental (d = 1.57) areas, surpasses that found in similar high-income studies using caregiver interventions, which have shown an average effect size between d = 0.3 to d = 0.6 [10,11]. It might be due to the more severe degree of impairment at baseline in this population, the community-based and relational model used by the Friendship Bench intervention, or the unique sensitivity of the social and environmental aspects of QoL assessments in this culture.

### 4.7 Strengths and Limitations

Strengths of this study include the use of prospective designs with matched pre-and post-intervention pairs, application of a scientifically sound and internationally validated QoL assessment tool with proven cross-cultural validity among sub-Saharan African populations [17,20], and reporting of statistically significant results along with the magnitude of changes. A multisite study conducted in two different geographical and hospital environments; Douala and Bertoua, enhances the transferability of findings within the country. This study captures the spillover effects of a task-shifted intervention strategy, which can be scaled up effectively in resource-poor settings.

A few limitations should be acknowledged. The fact that the sample size for the main paired sample was just 29 matches pair, even if sufficiently large to estimate large effects using this technique, significantly reduces the power of regression and correlation analyses that were performed as secondary analyses in the study. Attrition from 34 to 29 matched pairs may introduce selection bias if these participants differed systematically from completers.

## 5. Conclusion

Families of RTI survivors in Cameroon benefited significantly from quality of life improvement in all WHOQOL-BREF domains after their family member was exposed to the Friendship Bench psychological intervention, even though the family members did not undergo any intervention themselves. The magnitude of change was substantial to very large, uniform in terms of all domains, and was observed in both research sites. It should be noted that, despite being the most affected before the intervention, the social domain had the highest change value. Social relationship moderates the outcome, with the best performance belonging to children of the patients and poorer performance for the parents and extended family of the patient. Weak correlation between the patient PHQ-8 score and the family quality of life change may indicate a complex mechanism of spillover effects.

The current findings constitute new evidence of spillover effects arising from the implementation of task-shifting approaches to psychosocial care at the community level in a LMIC context in sub-Saharan Africa. The implications for practice include the incorporation of assessments of family members’ outcomes in the evaluative processes of psychosocial treatments, like the Friendship Bench. The findings point to the capacity of a patient-centered approach not only to positively affect patients but also the entire household as an interconnected system. Further studies are recommended using experimental designs to determine causality and investigate the mechanisms behind the positive household-level impact of patient-centered interventions.

## Data Availability

All data produced in the present study are available upon reasonable request to the authors.

## Declarations

The authors declare no competing interests. Research reported in this publication was supported by the Fogarty International Center of the National Institutes of Health (NIH) under award number U54W012087. This research is also supported by D-SINE Africa Seed Grant Program.

### Authors’ contributions

ADT conceptualised the study, led data collection and drafted the manuscript. FNDD, GMP and SM contributed to study design and supervision. RO, CJ and ACM provided critical revision of the manuscript. All authors read, reviewed, and approved the final version.

## Acknowledgements

The authors thank the patients and all family member participants who gave their time and shared their experiences.

## Notes

### Competing Interest Statement

The authors have declared no competing interest.

### Author Declarations

Ethical approval was obtained from the University of Cape Town Human Research Committee (HREC REF: 611/2025) and the University of Buea Institutional Review Board (2025: 2003- 04/UB/SG/IRB/FHS).

### Funding

- D-SINE Africa Pilot Project Seed Grants

## References

1. Global status report on road safety 2023. [visited on 11 June 2026]. Available: https://www.who.int/publications/i/item/9789240086517

2. Hung YW, Gallo JJ, Tol W, Syokau R, Bachani AM. Distress and resilience among unintentional injuries survivors in Kenya: A qualitative study. Rehabil Psychol. 2020;65: 45–53. doi:10.1037/rep0000289

3. Papadakaki M, Ferraro OE, Orsi C, Otte D, Tzamalouka G, von-der-Geest M, et al. Psychological distress and physical disability in patients sustaining severe injuries in road traffic crashes: Results from a one-year cohort study from three European countries. Injury. 2017;48: 297–306. doi:10.1016/j.injury.2016.11.011

4. Craig A, Tran Y, Guest R, Gopinath B, Jagnoor J, Bryant RA, et al. Psychological impact of injuries sustained in motor vehicle crashes: systematic review and meta-analysis. BMJ Open. 2016;6: e011993. doi:10.1136/bmjopen-2016-011993

5. Pozzato I, Tran Y, Gopinath B, Cameron ID, Craig A. The contribution of pre-injury vulnerability to risk of psychiatric morbidity in adults injured in a road traffic crash: Comparisons with non-injury controls. Journal of Psychiatric Research. 2021;140: 77–86. doi:10.1016/j.jpsychires.2021.05.064

6. Gopinath B, Jagnoor J, Kifley A, Dinh M, Craig A, Cameron ID. Predictors of health-related quality of life after non-catastrophic injury sustained in a road traffic crash. Annals of Physical and Rehabilitation Medicine. 2020;63: 280–287. doi:10.1016/j.rehab.2019.10.001

7. Hung KKC, Kifley A, Brown K, Jagnoor J, Craig A, Gabbe B, et al. PSYCHOLOGICAL DISTRESS, PAIN AND INSURANCE CLAIMS NEGATIVELY AFFECT LONG-TERM HEALTH-RELATED QUALITY OF LIFE AFTER ROAD TRAFFIC INJURIES. J Rehabil Med. 2022;54: 30. doi:10.2340/jrm.v54.30

8. Botta HW, Raykateeraroj N, Suh J, Lee D-K, Weinberg L. Outcomes of Older Adults Admitted to the ICU Following Trauma: A Scoping Review Identifying the Nonagenarian Evidence Gap. Cureus. 18: e102425. doi:10.7759/cureus.102425

9. Pozzato I, Tran Y, Arora M, McBain C, Middleton JW, Cameron ID, et al. Cumulative health burden and adjustment challenges following road traffic injuries: a controlled prospective study. BMC Med. 2025;23: 384. doi:10.1186/s12916-025-04084-0

10. Marshall CA, Nalder E, Colquhoun H, Lenton E, Hansen M, Dawson D, et al. Interventions to Address Burden Among Family Caregivers of Persons Aging with TBI: A Scoping Review. Brain Injury. 2018;28. doi:10.1080/02699052.2018.1553308

11. Sodders MD, Killien EY, Stansbury LG, Vavilala MS, Moore M. Race/Ethnicity and Informal Caregiver Burden After Traumatic Brain Injury: A Scoping Study. Health Equity. 2020;4: 304–315. doi:10.1089/heq.2020.0007

12. Agarwal TM, Muneer M, Asim M, Awad M, Afzal Y, Al-Thani H, et al. Psychological trauma in different mechanisms of traumatic injury: A hospital-based cross-sectional study. PLoS One. 2020;15: e0242849. doi:10.1371/journal.pone.0242849

13. Milandeep, Sagar R, Sagar S, Priyadarshini P, Kumar A, Alam J, et al. Impact of Psychosocial intervention on Quality of life in patients with post-traumatic limb amputation/s: a randomized controlled trial: Psychosocial care in post-traumatic amputees. Injury. 2026;57. doi:10.1016/j.injury.2026.113027

14. Psychological Distress following Injury in a Large Cohort of Thai Adults | PLOS One. [cited 11 June 2026]. Available: https://journals.plos.org/plosone/article?id=10.1371/journal.pone.0164767

15. Chibanda D, Bowers T, Verhey R, Rusakaniko S, Abas M, Weiss HA, et al. The Friendship Bench programme: a cluster randomised controlled trial of a brief psychological intervention for common mental disorders delivered by lay health workers in Zimbabwe. Int J Ment Health Syst. 2015;9: 21. doi:10.1186/s13033-015-0013-y

16. Effect of a Primary Care–Based Psychological Intervention on Symptoms of Common Mental Disorders in Zimbabwe: A Randomized Clinical Trial | Psychiatry and Behavioral Health | JAMA | JAMA Network. [cited 11 June 2026]. Available: https://jamanetwork.com/journals/jama/fullarticle/2594719

17. WHOQOL-BREF : introduction, administration, scoring and generic version of the assessment : field trial version, December 1996. [cited 11 June 2026]. Available: https://iris.who.int/items/8c5cd31a-77df-4d46-ad55-fc449fafe431

18. Skevington SM, Lotfy M, O’Connell KA. The World Health Organization’s WHOQOL-BREF quality of life assessment: Psychometric properties and results of the international field trial. A Report from the WHOQOL Group. Qual Life Res. 2004;13: 299–310. doi:10.1023/B:QURE.0000018486.91360.00

19. Heathcote K, Wullschleger M, Gardiner B, Morgan G, Barbagello H, Sun J. The Importance of Place of Residence on Hospitalized Outcomes for Severely Injured Trauma Patients: A Trauma Registry Analysis. The Journal of Rural Health. 2020;36: 381–393. doi:10.1111/jrh.12407

20. Gopinath B, Jagnoor J, Kifley A, Pozzato I, Dinh M, Craig A, et al. Twelve-month health outcomes for bicyclists and car occupants after a non-catastrophic traffic crash injury. Annals of Physical and Rehabilitation Medicine. 2021;64: 101368. doi:10.1016/j.rehab.2020.02.007

